# Developmental progression of the nasopharyngeal microbiome during childhood and association with the lower airway microbiome

**DOI:** 10.1101/2023.09.18.23295747

**Authors:** Ariel J. Hernandez-Leyva, Anne L. Rosen, Christopher P. Tomera, Elaina E. Lin, Elikplim H. Akaho, Allison M. Blatz, William R. Otto, Joey Logan, Lisa R. Young, Rebecca M. Harris, Andrew L. Kau, Audrey R. Odom John

**Affiliations:** Division of Allergy and Immunology, Department of Medicine and Center for Women’s Infectious Disease Research, Washington University School of Medicine, St. Louis, MO, 63110, USA; Perelman School of Medicine, University of Pennsylvania, Philadelphia, PA, USA; Department of Anesthesiology and Critical Care Medicine, Children’s Hospital of Philadelphia, Philadelphia PA; Division of Infectious Diseases, Department of Pediatrics, Children’s Hospital of Philadelphia, Philadelphia PA; Department of Medicine, John H. Stroger, Jr. Hospital of Cook County; Division of Critical Care Medicine, Department of Pediatrics, Nemours Children’s Hospital, Wilmington DE; Division of Infectious Disease, Cincinnati Children’s Hospital Medical Center; Department of Pediatrics, University of Cincinnati College of Medicine, Cincinnati Children’s Hospital Medical Center, Cincinnati, Ohio; Department of Biomedical and Health Informatics, Children’s Hospital of Philadelphia, Philadelphia PA; Division of Pulmonary and Sleep Medicine, Department of Pediatrics, Children’s Hospital of Philadelphia, Philadelphia PA; Department of Pathology and Laboratory Medicine, Children’s Hospital of Philadelphia, Philadelphia PA

## Abstract

**Background:** The upper (URT) and lower (LRT) respiratory tract feature distinct environments and responses affecting microbial colonization but investigating the relationship between them is technically challenging. We aimed to identify relationships between taxa colonizing the URT and LRT and explore their relationship with development during childhood.

**Methods:** We employed V4 16S rDNA sequencing to profile nasopharyngeal swabs and tracheal aspirates collected from 183 subjects between 20 weeks and 18 years of age. These samples were collected prior to elective procedures at the Children’s Hospital of Philadelphia over the course of 20 weeks in 2020, from otherwise healthy subjects enrolled in a study investigating potential reservoirs of SARS-CoV-2.

**Findings:** After extraction, sequencing, and quality control, we studied the remaining 124 nasopharyngeal swabs and 98 tracheal aspirates, including 85 subject-matched pairs of samples. V4 16S rDNA sequencing revealed that the nasopharynx is colonized by few, highly-abundant taxa, while the tracheal aspirates feature a diverse assembly of microbes. While no taxa co-occur in the URT and LRT of the same subject, clusters of microbiomes in the URT correlate with clusters of microbiomes in the LRT. The clusters identified in the URT correlate with subject age across childhood development.

**Interpretations:** The correlation between clusters of taxa across sites may suggest a mutual influence from either a third site, such as the oropharynx, or host-extrinsic, environmental features. The identification of a pattern of upper respiratory microbiota development across the first 18 years of life suggests that the patterns observed in early childhood may extend beyond the early life window.

**Funding:** Research reported in this publication was supported by NIH T32 GM007200 (AJH), F30 DK127584 (AJH), NIH/NIAID R21AI154370 (AOJ, ALK), NIH/NICHD R01HD109963 (AOJ, ALK), and NIH/NICHD R33HD105594 (AOJ). Dr. John is an Investigator in the Pathogenesis of Infectious Diseases of the Burroughs Welcome Fund.

## Introduction

The microbial communities of the airway are an important regulator of human respiratory health that develop alongside the host and can experience dysbiosis in disease. For example, colonization with specific health-associated, commensal microbes has been linked to reduced inflammation during *Pseudomonas aeruginosa* pneumonia in patients with cystic fibrosis.^1^ On the other hand, viral respiratory infections in young children lead to a preponderance of upper airway *Moraxella*, *Haemophilus*, and *Streptococcus* species that are associated with increased risk for chronic wheeze and later development of asthma.^2,3^

The composition of the airway microbiota, and thus its influence on host health, is shaped by both host-intrinsic and external factors.^4^ Host-intrinsic factors such as pH, surface area, and partial pressure of gases vary throughout the respiratory tract and form distinct environments that foster diverse microbial organisms. External factors include birth mode and feeding during infancy, home and school/daycare environment during childhood, and exposure to smoking, antibiotics, and infections throughout adulthood.^4^ These factors play an important role in determining how the microbiota is seeded and cultivated at key developmental life stages. Additionally, features such as season, temperature, and humidity continue to shape rapid fluctuations in community composition.^5^ Understanding how these factors play a role in defining the healthy airway microbiota is important to identify deviations towards dysbiosis.

The respiratory tract is conceptually divided into two regions with mucosal surfaces superior to the larynx (including the nares, nasal vault, oronasopharynx, and laryngopharynx) comprising the upper respiratory tract (URT) and those inferior to the larynx (including the trachea, bronchi, and small airways extending to alveoli) comprising the lower respiratory tract (LRT).^6^ Despite anatomical continuity, the upper and lower respiratory tracts exhibit differences in the epithelial cell populations, immunological barriers, and environmental phenomena that support distinct ecologies. For example, the microbial communities of the URT colonize in distinct spatial patterns across an otherwise contiguous mucosa in response to the different epithelia and host-intrinsic features.^7,8^ Thus, the primary determinant of URT community composition is fitness to the environmental niche.^9^ On the other hand, the composition of LRT microbial communities is less dependent on biogeography, and the same taxa can be detected throughout, although the most ecologically rich community belongs to the upper trachea.^10^ Microbial community richness decreases proportional to anatomical depth into the LRT, supporting a model where immigration of taxa by aspiration and emigration of taxa through mucociliary clearance, cough, or phagocytosis by innate immune cells are the primary factors shaping community composition, rather than regional niche differences..^11,12^ Previous work has found that the microbial composition of the oropharynx was strongly associated with that of the LRT microbiota^13^. However, the influence of the various other sites of the upper airway on lower airway composition has not been well catalogued, especially during pediatric development.

In this study, we looked at nasopharyngeal swabs and tracheal aspirates samples from 183 pediatric subjects collected during elective outpatient surgical procedures. Samples were profiled using V4 16S rDNA sequencing to explore the relationship between the host and the microbiomes of the upper and lower respiratory tract. These samples were initially collected as part of a study to determine the degree of concordance in the results of reverse-transcriptase polymerase chain reaction assays for SARS-CoV-2 between upper and lower respiratory tracts.^14,15^

## Methods

### Study Cohort

The subject cohort was a fraction of SARS-CoV-2 RT-PCR-negative subjects from a convenience sample of pediatric subjects under the age of 18 undergoing elective surgical interventions at the Children’s Hospital of Philadelphia as previously described.^14,15^ The study was reviewed by the Institutional Review Board at Children’s Hospital of Philadelphia and consent was obtained from the patients’ guardians. Subjects were enrolled between July 10^th^ and November 24^th^, 2020. Tracheal aspirate samples were collected under general anesthesia by either the anesthesiologist or pulmonologist at the time of procedure and nasopharyngeal swabs were collected the same day. Samples were stored at -20°C until shipment. Between 10 μL to 1 mL of each sample or negative controls were shipped on dry ice to Washington University in St. Louis, where they were stored at -80°C until processing.

### Sample Processing and V4 16S rDNA Sequencing

All available volume of the tracheal aspirates and nasopharyngeal swabs of 183 subjects were extracted using the QIAGEN Microbiome DNA Extraction Kit (QIAGEN CAT#51704) following manufacturer’s directions. Extraction was completed in batches of 20-22 samples with at least one negative control in every batch. Negative controls included PBS used in extractions or sterile transport buffer used for tracheal aspirates. After extraction, V4 – V9 16S rDNA sequences were initially amplified over 20 cycles of PCR using primers 515F and 1492R. Subsequently, the V4 16S rDNA region was amplified 30 cycles from pre-amplified DNA using barcoded variations of the 515F and 806R primers previously described in Hazan et al.^16^ Amplicons were sequenced using an Illumina MiSeq instrument with 2x250 bp chemistry. FASTQ files were demultiplexed and processed as described in Wilson et. al.^17^ The tables of ASVs for each sample type— nasopharyngeal swabs and tracheal aspirates—were then filtered using decontam v1.16.0.^18^ BLAST searches against the 16S ribosomal RNA sequences database were completed using the online BLASTn software.^19^ Samples remaining after decontamination and filtering are summarized in Table 1. The details of decontamination and filtering are presented in the supplementary information and Supplementary Figure S1.

**Table 1:** Summary of demographics for available subject samples.

|  | Nasopharyngeal Swab | Tracheal Aspirates | Matched |
| --- | --- | --- | --- |
| **N =** | 124 | 98 | 85 |
| **Age (years)** |  |  |  |
| *Mean (Standard Deviation)* | 7·51 (5·31) | 7·07 (5·18) | 7·16 (5·19) |
| *Median (Min - Max)* | 6·47 (0·56 - 17·86) | 5·57 (0·39 - 17·85) | 6·13 (0·56 - 17·85) |
| **Sex** |  |  |  |
| *Male* | 75 | 57 | 50 |
| *Female* | 49 | 41 | 35 |
| **Past Medical History** |  |  |  |
| *Any chronic respiratory disease* | 28 | 19 | 18 |
| *Any chronic disease* | 39 | 27 | 26 |
| *Born Premature* | 8 | 6 | 6 |
| **Race** |  |  |  |
| *White* | 75 | 54 | 50 |
| *Black or African American* | 23 | 21 | 18 |
| *Asian* | 5 | 3 | 3 |
| *American Indian/Alaska native* | 1 | 1 | 1 |
| *Multiple* | 2 | 2 | 2 |
| *Other* | 18 | 17 | 11 |
| **Ethnicity** |  |  |  |
| *Hispanic or Latino* | 18 | 14 | 11 |
| **Seasonality (Weeks from Study Start)** |  |  |  |
| *Mean (Standard Deviation)* | 13·2 (4·67) | 13·34 (4·56) | 13·41 (4·66) |
| *Median (Min - Max)* | 12 (4 - 20) | 12 (4 - 20) | 12 (4 - 20) |
| **Intervention** |  |  |  |
| *General Procedure* | 17 | 11 | 9 |
| *Ear/Nose/Throat Procedure* | 39 | 34 | 30 |
| *Urologic Procedure* | 11 | 13 | 10 |
| *Orthopedic Procedure* | 8 | 6 | 5 |
| *NeuroProcedure* | 9 | 8 | 6 |
| *Dental Procedure* | 13 | 10 | 10 |
| *Pulmonary Procedure\** | 8 | 0 | 0 |
| *Gastrointestinal Endoscopy\** | 7 | 0 | 0 |
| *Plastics Procedure* | 5 | 4 | 3 |
| *Oral and Maxillofacial Procedure* | 3 | 3 | 3 |

*Asterisks (\*) indicate statistically significant difference between tracheal aspirate samples and nasopharyngeal swabs*

### Statistical Analysis

Statistics and all data analyses were performed in R version 4.2.1. Data are represented as means with standard deviation unless otherwise specified. Tests conducted for statistical significance are described in the text and figure legends. Adjustment for p-values was performed using Benjamini-Hochberg correction when necessary. Violin plots and box plots display the IQR and the whiskers display 1.5* the IQR. The following symbols are used to denote significance: * p <0·05, ** p <0·01, *** p < 0·001, **** p < 0·0001. Ecological analysis of V4 16S rDNA sequencing data was completed as previously described in Wilson et. al.^17^ Dirichlet multinomial distributions were modeled using the DirichletMultinomial v1.38.0 and the microbiome v1.18.0 packages in R.

## Results

### Description of Cohort

The samples from 183 pediatric surgical patients recruited from the Children’s Hospital of Pennsylvania were included in the study. The mean age of subjects was 7·79 years (SD = 5·35) with a range from 5 months to 18 years, of which 112 were male and 71 were female. Twelve different procedure types were identified, with the most common being ear-nose-throat procedures representing 32·2% of subjects. All samples from subjects were collected during a 20-week span of time (July 10^th^, 2022 – November 24^th^, 2022). At the time, the Philadelphia, PA metro area was under COVID-19 mitigation, including masking and distancing, and the incidence of common respiratory viral infections was low.

### Overview of the V4 16S rDNA profiles from nasopharyngeal swabs and tracheal aspirates of pediatric patients

To review the results of V4 16S rDNA profiling we visualized the data both summarized to the phyla level, and at the level of individual ASVs. Both the URT and LRT microbiota are occupied by six major phyla: Bacillota, Actinomycetota, Pseudomonadota, Bacteriodota, Fusobacteriota, and Verrucomicrobiota (Figure 1A, B). The most common phylum in both types of samples is Bacillota; however, the second most common phylum varies between the nasopharyngeal swabs and the tracheal aspirates. In the nasopharyngeal swab samples, ASVs are more frequently classified as Actinomycetota, whereas in tracheal aspirate samples ASVs are more frequently classified as Pseudomonadota. These overarching differences in community structure are apparent by beta-diversity analysis, where nasopharyngeal swab microbiomes cluster separately from tracheal aspirate microbiomes (Figure 1C, adonis2 R^2^: 0·154, p-value < 0·0002). Examining the composition of the nasopharyngeal microbiomes more closely, we observe that most samples carry either the Bacillota *Dolosigranulum pigrum,* or *Staphylococcus aureus* (Figure 1D). The next most common taxa tend to be the Actinomycetota *Corynebacterium,* of which we were only able to classify one to the species level: *Corynebacterium macginleyi.* Among the most abundant taxa identified in tracheal aspirate microbiomes are the Pseudomonadota *Escherichia coli* and the Bacillota *Gamella hemolysans*, *Dolosigranulum pigrum, Streptococcus*, and *Staphylococcus* (Figure 1E). *E. coli* is an unexpected community member of the airway microbiome. For this reason, we investigated the sequence of the ASV using BLAST and discovered that the same sequence shared 100% identity with the V4 16S rDNA sequences of several Enterobacteriaceae including *Shigella* species, *Escherichia* species, and *Pseudescherichia vulneris*. While our ability to resolve the taxonomic composition is limited by marker gene sequencing, our data highlights marked differences between the microbiomes of the nasopharyngeal swabs and tracheal aspirates. To further resolve the microbiomes of the nasopharyngeal swabs and tracheal aspirates, we conducted a co-occurrence analysis to identify sets of taxa within each community that may share an ecological relationship. We visualized this analysis as networks within the nasopharyngeal swab microbiomes (Figure 2A) and tracheal aspirate microbiomes (Figure 2B). In the nasopharyngeal microbiomes, we identified a few relationships among the most abundant taxa including a positive co-occurrence between *D. pigrum* and *Corynebacterium* species that has previously been reported in the literature.^20^ Notably, both species also negatively co-occur with *S. aureus*, another phenomenon that has been previously described.^20^ There is a greater number of co-occurring taxa in the tracheal aspirate microbiomes. A cluster of three taxa, *Bacillus simplex*, *Lactobacillus mali*, *and Paracoccus aminovorans* sits apart from the main body of positive co-occurring taxa that includes various ASVs attributed to *Streptococcus*. Previously, lower airway *Streptococcus* colonization has been linked to aspiration or microaspiration of microorganisms colonizing the oropharynx.^11,21^ *Bacillus* species and *Paracoccus* species have been well described in soil, although some species have acted as opportunistic pathogens in humans.^22–24^ *Lactobacillus mali* has previously been explored as a possible probiotic affiliated with improved airway health in the context of infection, cystic fibrosis, and asthma.^25^ Airway colonization with *Lactobacillus* has been ascribed to oropharyngeal reflux.^26,27^

**Figure 1:**
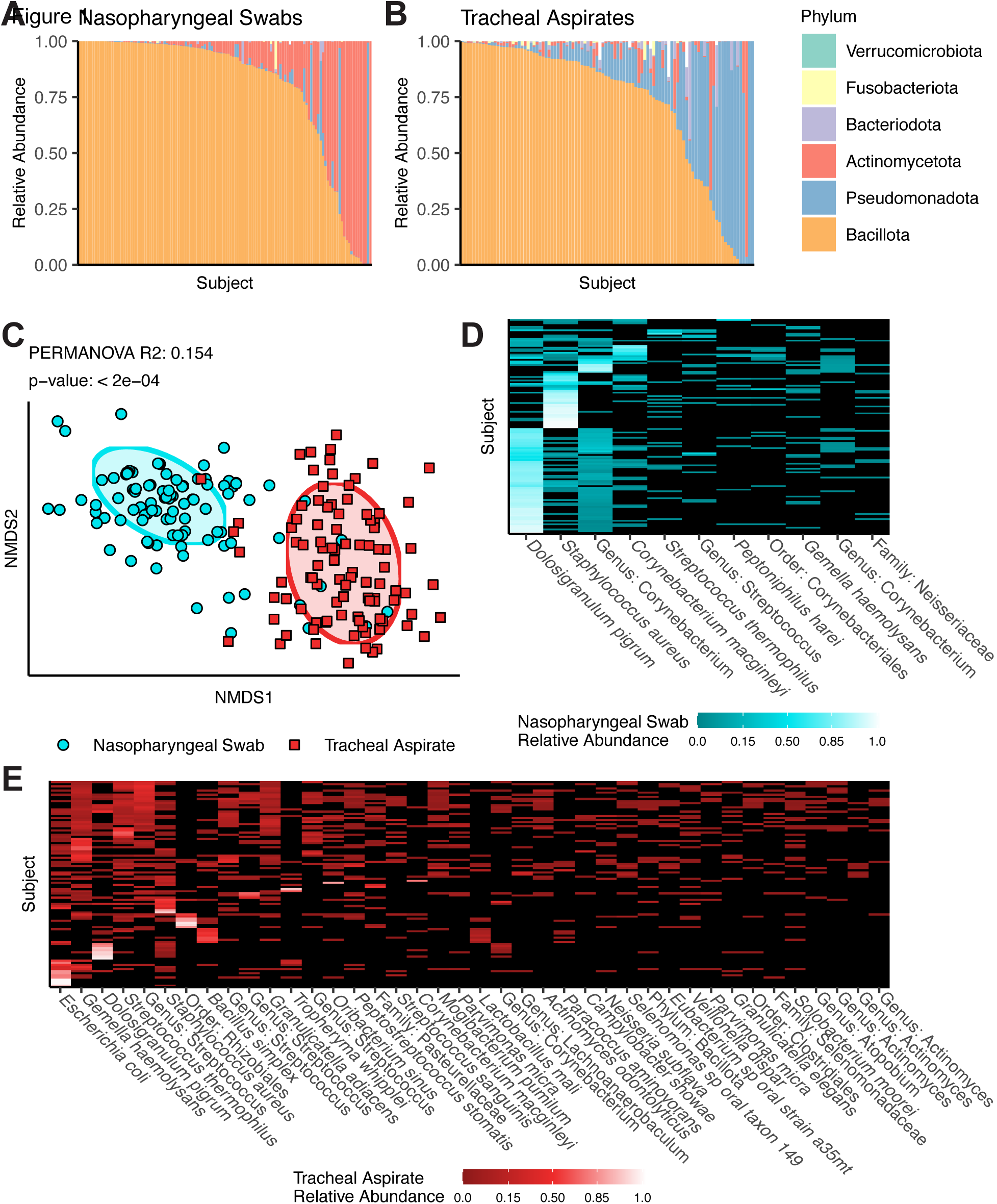
Summary of V4 16S rDNA Sequencing results from upper and lower airway samples. A - B) Taxonomic relative abundance means plots depicting phyla level abundances for each sample in either the nasopharyngeal swabs (A, N = 124) or tracheal aspirates (B, N = 98). White bars reflect the proportion of taxa that were not classified to the phyla level. C) Non-metric dimensional scaling depicting the Unifrac distances between all samples. Results of PERMANOVA reflect the difference between nasopharyngeal swab and tracheal aspirate microbiomes. D) Heatmap reflecting taxa detected in at least 10% the nasopharyngeal swabs samples. E) Heatmap reflecting taxa detected in at least 10% of the tracheal aspirate samples.

**Figure 2:**
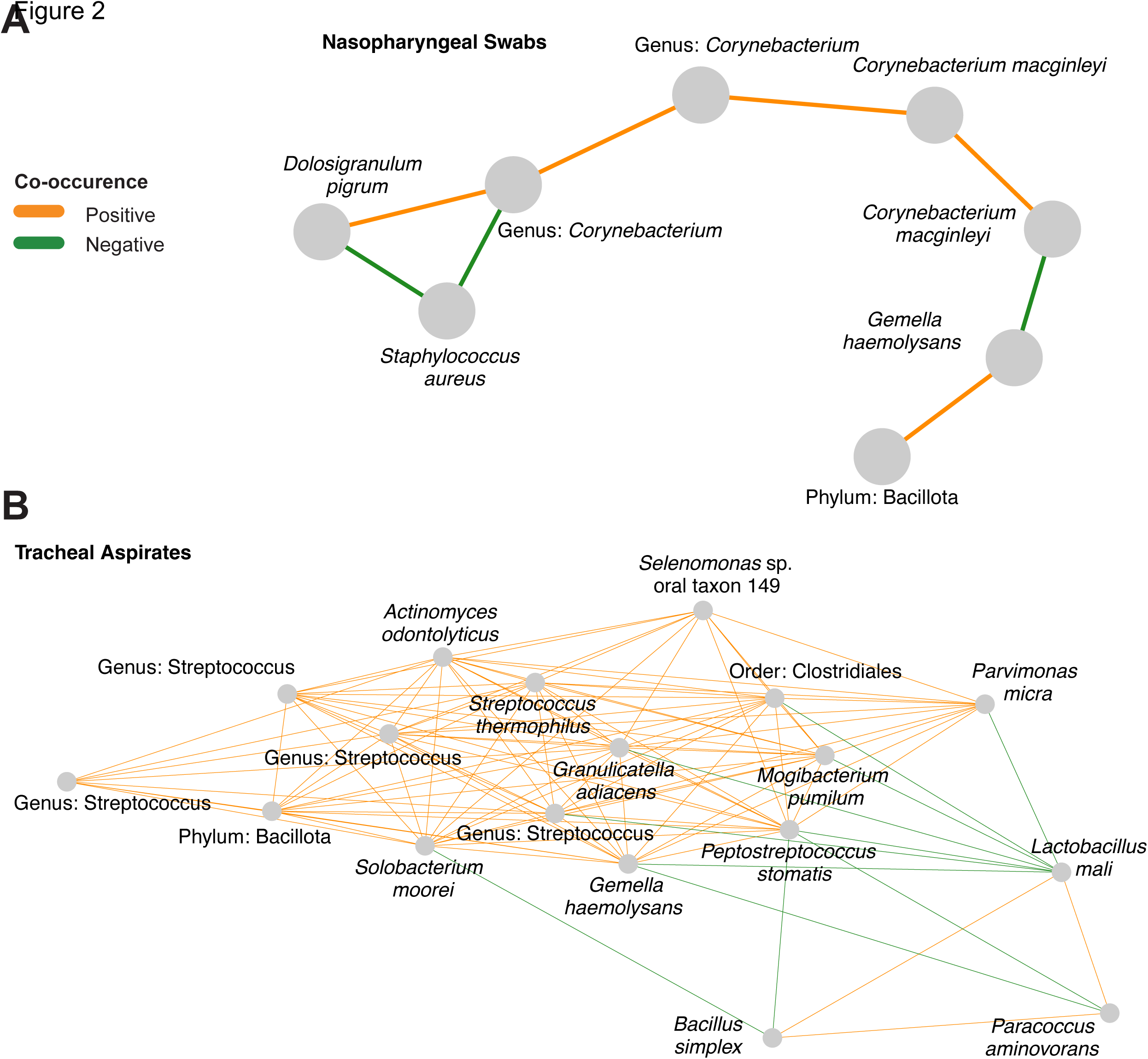
Co-occurrence networks reveal interactions in nasopharyngeal swabs and tracheal aspirate microbiomes. A) Co-occurrence network generated from V4 16S rDNA profiling the nasopharyngeal samples. Nodes were only included if they shared an edge with a minimum effect size of 0.05. B) Co-occurrence network generated from V4 16S rDNA profiling the tracheal aspirate samples. Nodes were only included if they shared an edge with a minimum effect of 0.10.

### Defined clusters of taxa describe the upper and lower respiratory tract microbiomes

Since we observed several defined networks of taxa within the co-occurrence analysis of the respiratory tract microbiomes, we thought that the nasopharyngeal and tracheal microbiomes might be well described by clusters of these taxa. We employed Dirichlet multinomial modeling to infer clusters from the nasopharyngeal and tracheal microbiomes separately. We found that the nasopharyngeal microbiomes grouped best into two distinct clusters. However, three clusters were nearly as optimal and demonstrated more similarities with the co-occurrence network (Figure 3A). The cluster NS1 was highly colonized by *D. pigrum* and *Corynebacterium* (Supplementary Figure S2A). Subjects assigned NS2 were more likely to be colonized by a mixture of microbes, including *D. pigrum, S. aureus*, and *Corynebacterium,* but also featured a greater fraction of subjects colonized with *Streptococcus* spp. All subjects in the final cluster, NS3, were colonized with *S. aureus*, and featured a much lower rate of colonization with *D. pigrum* or *Corynebacterium* than the other clusters. Overall, we found that the clusters explained a significant proportion of the variation in alpha diversity between subjects (Figure 3B, ANOVA p-value < 0·0001), with NS2 displaying the greatest alpha diversity on average. The clusters also explained a significant proportion of the variation in the beta-diversity between nasopharyngeal microbiomes (Figure 3C, adonis2 R^2^: 0·191, p-value: 0·0002), supporting the idea that the Dirichlet multinomial modeling discovered important differences.

**Figure 3:**
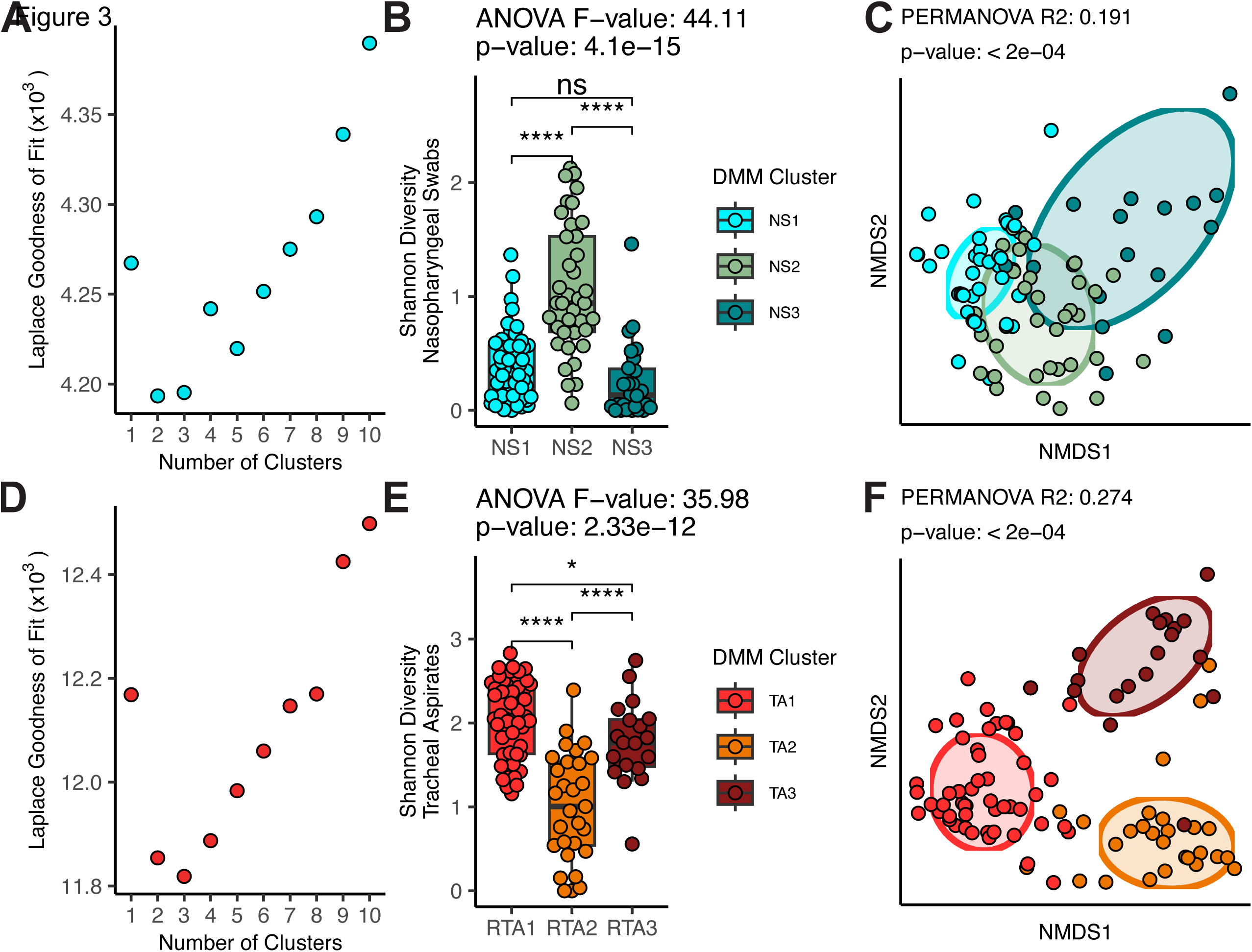
Summary of clusters identified by Dirichlet multinomial modeling of the nasopharyngeal swab microbiomes and tracheal aspirate microbiomes. A) Laplace goodness of fit comparison for models produced with various numbers of clusters for the nasopharyngeal swabs. B) A comparison of Shannon diversity measures for the microbiota of the nasopharyngeal swabs separated by assigned cluster. Nasopharyngeal swabs are divided into three clusters: NS1 (N = 60), NS2 (N = 38), and NS3 (N = 26). C) Non-metric dimensional scaling analysis of the unweighted Unifrac distances between nasopharyngeal microbiomes. D) Laplace goodness of fit comparison for models produced with various numbers of clusters for the tracheal aspirates. E) A comparison of Shannon diversity measures for the microbiota of the tracheal aspirates separated by assigned cluster. Tracheal aspirates are divided into three clusters: TA1 (N = 50), TA2 (N = 29), and TA3 (N = 19). F) Non-metric dimensional scaling analysis of the unweighted Unifrac distances between tracheal aspirate microbiomes. ANOVA was used to test whether the clusters explained a significant proportion of variance in the alpha diversity of subjects. T-tests with Benjamini-Hochberg correction was used to demonstrate between-cluster differences. NMDS analyses of beta diversity were rotated to display axes with the greatest variation between clusters. PERMANOVA was conducted to test whether the assigned clusters explained a significant proportion of the variance in the data.

We also found that our tracheal aspirate microbiomes grouped best into three (Figure 3D). TA1, featured high rates of colonization with many different taxa, including *Streptococcus, Gamella hemolysans, S. aureus* as well as *Mogibacterium pumilum, Actinomyces odontolyticus,* and *Solobacterium moorei* (Supplementary Figure 2B). TA2, predominantly featured high rates of colonization with the ASV previously identified as *Escherichia coli*. TA3 featured high rates of colonization with *Bacillus simplex, Lactobacillus malii,* and *Paracoccus aminovorans*. As expected, several of these clusters represent features observed in the co-occurrence networks, including the relationship between *D. pigrum, S. aureus,* and *Corynebacterium* observed in the upper airway. As among the nasopharyngeal microbiomes, we also found that the tracheal aspirate clusters explained a significant proportion of the variance in alpha diversity, (Figure 3E, ANOVA p-value < 0·0001) with TA1 featuring the greatest alpha diversity on average, and TA2 featuring the least. The assigned clusters also explained a significant proportion of the variance in the beta-diversity between the tracheal aspirate microbiomes (Figure 3F, adonis2 R^2^: 0·274, p-value < 0·0002).

We evaluated whether any of the clusters were associated with subject demographics. We observed a strong correlation between the nasopharyngeal clusters and age (Table 2, Supplementary Table 1, ANOVA p-value: 0·0001), with NS1 representing a younger cohort on average than NS2 or NS3. There was an association between ethnicity and assigned tracheal aspirate cluster (ANOVA p-value = 0·022), with a greater proportion of subjects responding Hispanic or Latino among TA3. We also observed a slight enrichment of subjects undergoing orthopedic procedures in NS3, and only observed subjects undergoing oral and maxillofacial procedures in the NS2 group. TA2 meanwhile was enriched for subjects undergoing ear, nose, and throat procedures (Table 3, Supplementary Table 1).

**Table 2:** Summary of demographics for Dirchlet-multinomial defined nasopharyngeal clusters.

|  | NS1 | NS2 | NS3 |
| --- | --- | --- | --- |
| **N =** | 60 | 38 | 26 |
| **Age (years)\*\*\*** |  |  |  |
| *Mean (Standard Deviation)* | 5·47 (4·48) | 9·58 (6·08) | 9·19 (4·13) |
| *Median (Min - Max)* | 3·9 (0·56 - 17·83) | 10·41 (0·63 - 17·86) | 9·2 (0·97 - 17·45) |
| **Sex** |  |  |  |
| *Male* | 38 | 21 | 16 |
| *Female* | 22 | 17 | 10 |
| **Past Medical History** |  |  |  |
| *Any chronic respiratory disease* | 12 | 10 | 6 |
| *Any chronic disease* | 13 | 14 | 12 |
| *Born Premature* | 5 | 2 | 1 |
| **Race** |  |  |  |
| *White* | 35 | 21 | 19 |
| *Black or African American* | 10 | 8 | 5 |
| *Asian* | 3 | 2 | 0 |
| *American Indian/Alaska native* | 0 | 1 | 0 |
| *Multiple* | 2 | 0 | 0 |
| *Other* | 10 | 6 | 2 |
| **Ethnicity** |  |  |  |
| *Hispanic or Latino* | 12 | 5 | 1 |
| **Seasonality (Weeks from Study Start)** |  |  |  |
| *Mean (Standard Deviation)* | 13 (4·34) | 14·5 (4·99) | 11·77 (4·63) |
| *Median (Min - Max)* | 12 (4 - 20) | 17 (4 - 20) | 11 (4 - 20) |
| **Intervention** |  |  |  |
| *General Surgery* | 9 | 5 | 3 |
| *Ear/Nose/Throat Surgery* | 21 | 10 | 8 |
| *Urologic Surgery* | 8 | 3 | 0 |
| *Orthopedic Surgery\*\*\** | 1 | 2 | 5 |
| *Neurosurgery* | 5 | 2 | 2 |
| *Dental Surgery* | 8 | 2 | 3 |
| *Pulmonary Surgery* | 2 | 2 | 4 |
| *Gastrointestinal Endoscopy* | 2 | 3 | 2 |
| *Plastics Surgery* | 1 | 3 | 1 |
| *Oral and Maxillofacial Surgery\** | 0 | 3 | 0 |

**Table 3:**
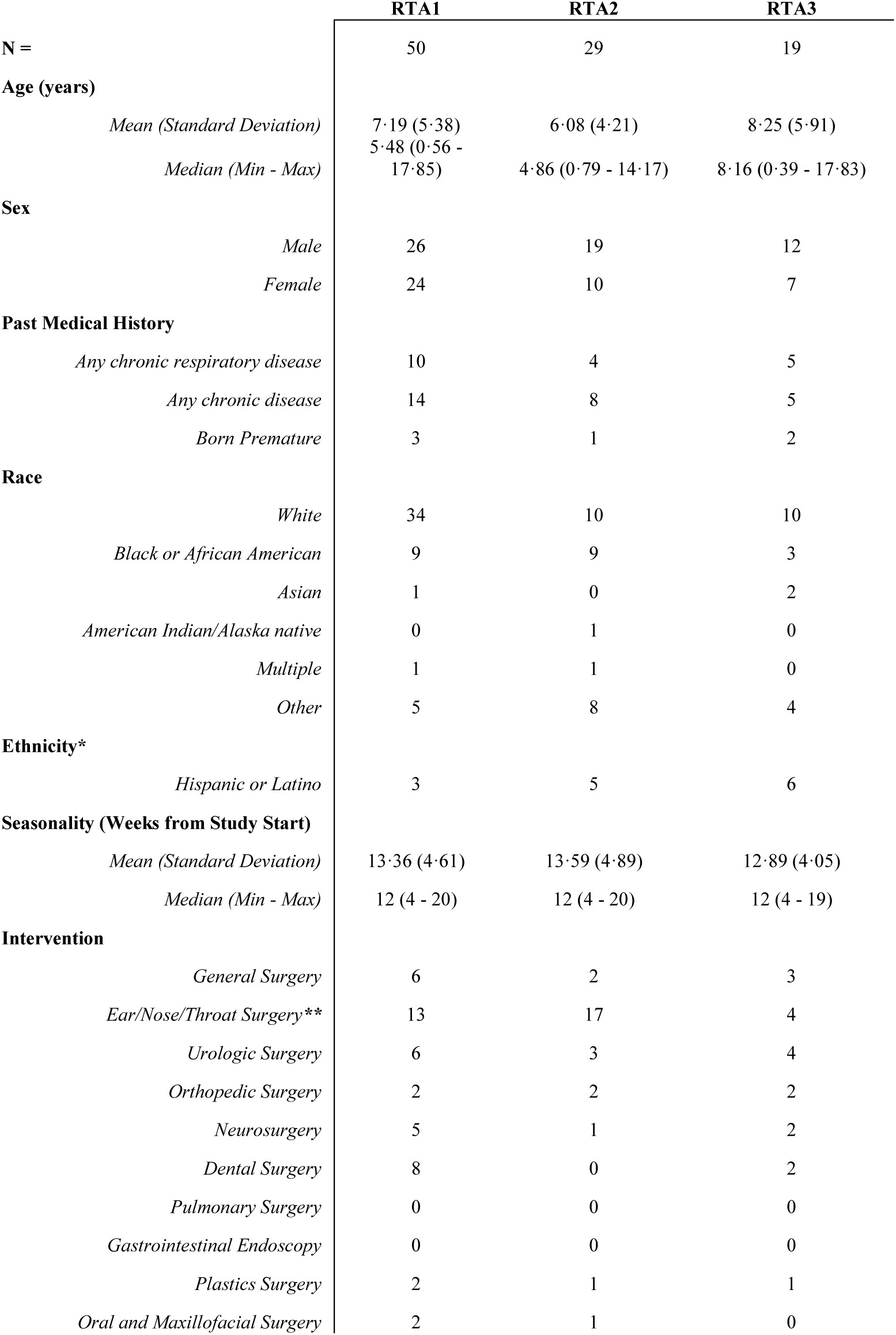
Summary of demographics for Dirchlet-multinomial defined tracheal apsirate clusters.

|  | RTA1 | RTA2 | RTA3 |
| --- | --- | --- | --- |
| **N =** | 50 | 29 | 19 |
| **Age (years)** |  |  |  |
| *Mean (Standard Deviation)* | 7·19 (5·38) | 6·08 (4·21) | 8·25 (5·91) |
| *Median (Min - Max)* | 5·48 (0·56 - 17·85) | 4·86 (0·79 - 14·17) | 8·16 (0·39 - 17·83) |
| **Sex** |  |  |  |
| *Male* | 26 | 19 | 12 |
| *Female* | 24 | 10 | 7 |
| **Past Medical History** |  |  |  |
| *Any chronic respiratory disease* | 10 | 4 | 5 |
| *Any chronic disease* | 14 | 8 | 5 |
| *Born Premature* | 3 | 1 | 2 |
| **Race** |  |  |  |
| *White* | 34 | 10 | 10 |
| *Black or African American* | 9 | 9 | 3 |
| *Asian* | 1 | 0 | 2 |
| *American Indian/Alaska native* | 0 | 1 | 0 |
| *Multiple* | 1 | 1 | 0 |
| *Other* | 5 | 8 | 4 |
| **Ethnicity\*** |  |  |  |
| *Hispanic or Latino* | 3 | 5 | 6 |
| **Seasonality (Weeks from Study Start)** |  |  |  |
| *Mean (Standard Deviation)* | 13·36 (4·61) | 13·59 (4·89) | 12·89 (4·05) |
| *Median (Min - Max)* | 12 (4 - 20) | 12 (4 - 20) | 12 (4 - 19) |
| **Intervention** |  |  |  |
| *General Surgery* | 6 | 2 | 3 |
| *Ear/Nose/Throat Surgery\*\** | 13 | 17 | 4 |
| *Urologic Surgery* | 6 | 3 | 4 |
| *Orthopedic Surgery* | 2 | 2 | 2 |
| *Neurosurgery* | 5 | 1 | 2 |
| *Dental Surgery* | 8 | 0 | 2 |
| *Pulmonary Surgery* | 0 | 0 | 0 |
| *Gastrointestinal Endoscopy* | 0 | 0 | 0 |
| *Plastics Surgery* | 2 | 1 | 1 |
| *Oral and Maxillofacial Surgery* | 2 | 1 | 0 |

### Diversity of the nasopharyngeal microbiome does not correlate with the diversity of the tracheal microbiome

We compared the microbial diversity of the nasopharyngeal swab microbiomes against the diversity of the tracheal aspirate microbiomes using Shannon’s diversity (Figure 4A). We observed that the tracheal aspirates seem to have a greater diversity on average than the nasopharyngeal swabs. Using the 85 matched samples, we measured the correlation in diversity between nasopharyngeal swabs and the tracheal aspirates and found a weak, non-significant association (Figure 4B, Pearson’s R^2^: 0·045, p-value: 0·052). We also evaluated whether taxa that could be found in both the nasopharyngeal swabs and tracheal aspirates might co-occur within the same subject. Once again, we leveraged the 85 subject-matched nasopharyngeal and tracheal samples but found that none of the fifteen taxa that appear in both anatomic sites significantly co-occurred in upper and lower airways of the same subject more than could be expected by chance alone (Table 4). We sought to identify whether the nasopharyngeal and the tracheal community type might share a relationship, so we also tested whether the clusters assigned to the upper airway communities were associated with the clusters assigned to the lower airway communities. We found that there was a significant association between the clusters of the nasopharyngeal swabs and the clusters of the tracheal aspirates, with the mixed cluster NS2 co-occurring with *Bacillus simplex* containing TA3 (Figure 4C).

**Figure 4:**
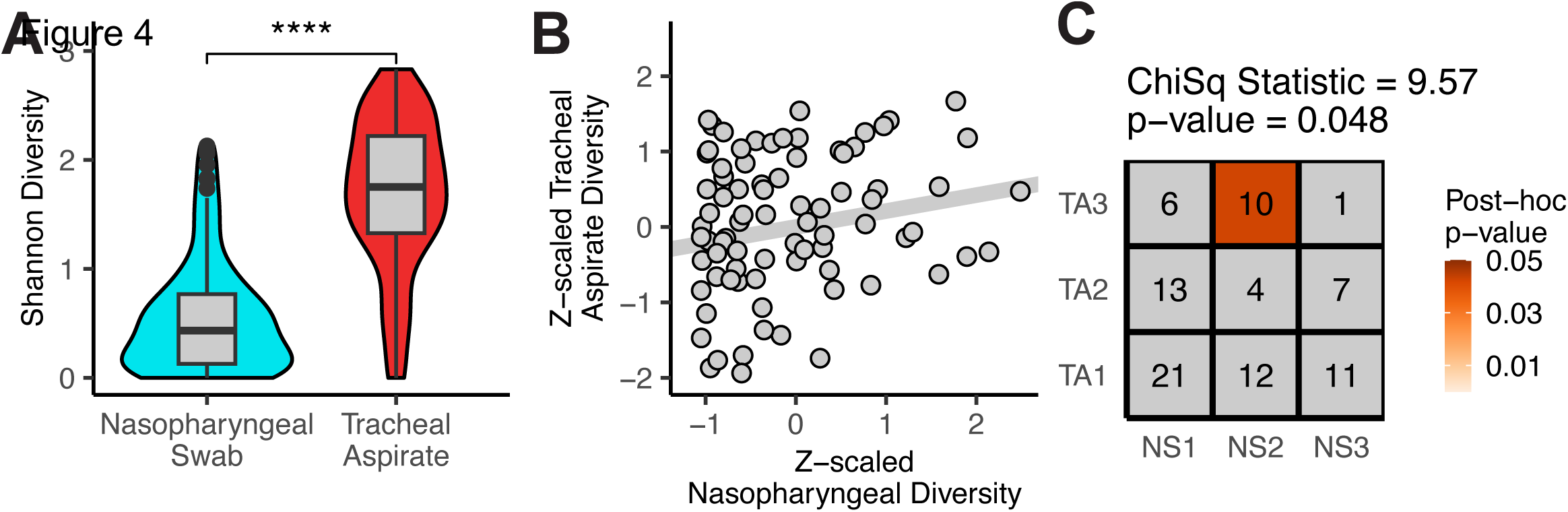
Correlating features of the nasopharyngeal swabs and tracheal aspirates. A) Shannon diversity comparison between nasopharyngeal swabs (N = 124) and tracheal aspirates (N = 98). Mean difference was assessed using Student’s t-test. B) Comparison of the correlation between tracheal aspirate diversity and nasopharyngeal diversity for the 85 paired samples. The strength of the relationship was measured with Pearson’s correlation. C) Chi-square test comparing clusters assigned to the nasopharyngeal swabs against clusters assigned to the tracheal aspirates. A post-hoc chi-square test was used to identify unlikely correlations.

**Table 4:**
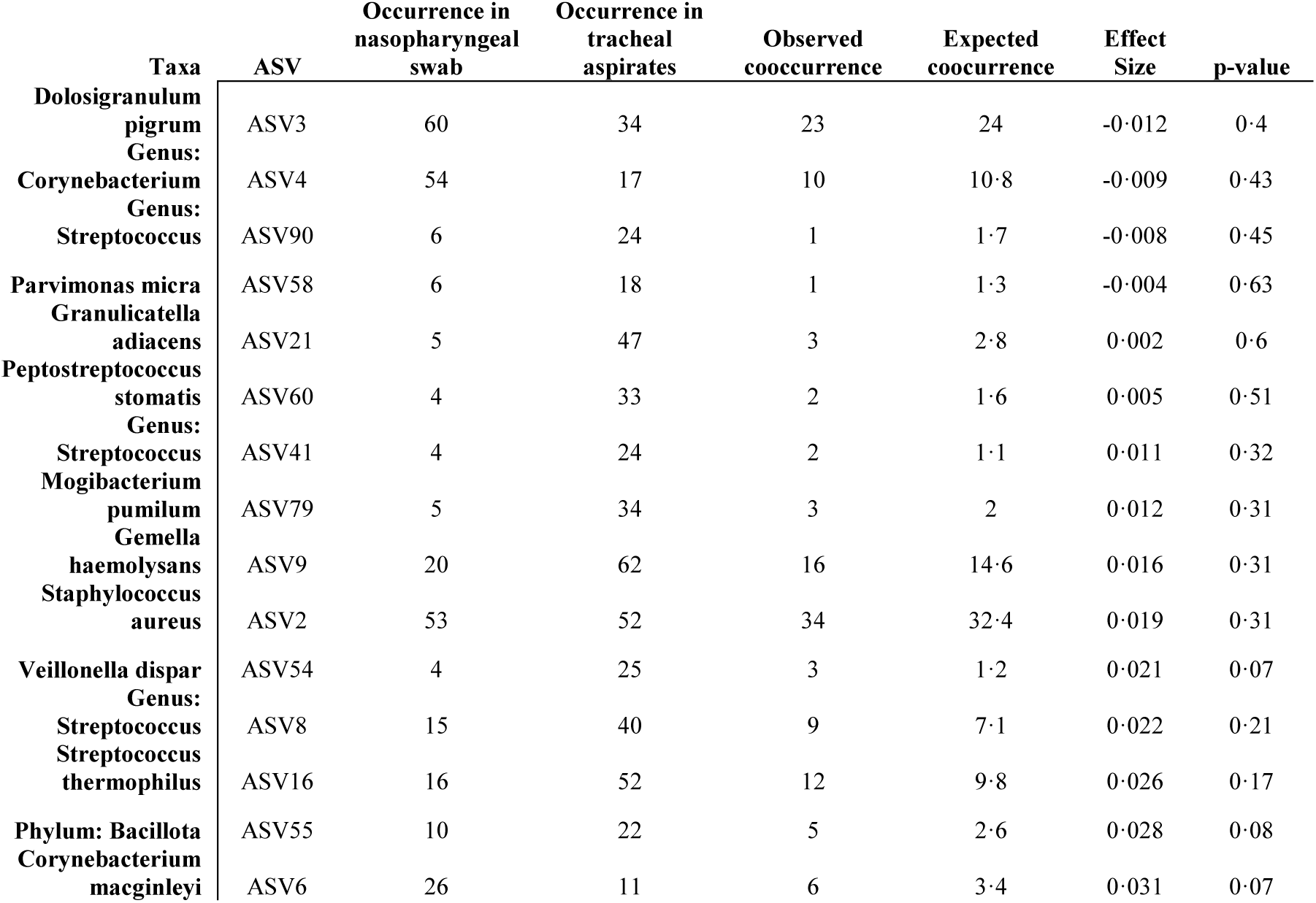
Summary of taxa co-occurring in the nasopharyngeal swab and tracheal aspirate of the same individual.

### Age influences the nasopharyngeal swab microbiome across childhood

We independently compared the nasopharyngeal swab microbiomes and tracheal aspirate microbiomes against demographic factors to identify important subject features. We first performed an ANOVA comparing the alpha diversity of nasopharyngeal swabs across all available demographic factors (Figure 5A). We found that subject age was the most important feature, but the date of sample collection, and whether the subject underwent oral/maxillofacial procedures, gastrointestinal/endoscopic procedures, or dental procedures all explained a significant proportion of the variance in alpha diversity. Further investigation demonstrated a weak but significant correlation between age and nasopharyngeal alpha diversity (Pearson’s R^2^: 0·053, p-value: 0·010, Figure 5B). The correlation between alpha diversity and seasonality was even weaker and not significant (Pearson’s R^2^: 0·025, p-value: 0·078, Supplementary Figure S3A). We also examined the procedure types identified as important by ANOVA but found a significant difference only in the mean alpha diversities between subjects undergoing oral/maxillofacial procedures and those that were not (Supplementary Figure S3B). We also investigated the influence of the same demographic factors on the Unifrac distances between nasopharyngeal swab microbiomes and found that age alone explained a significant proportion of the variance (Figure 5C-D).

**Figure 5:**
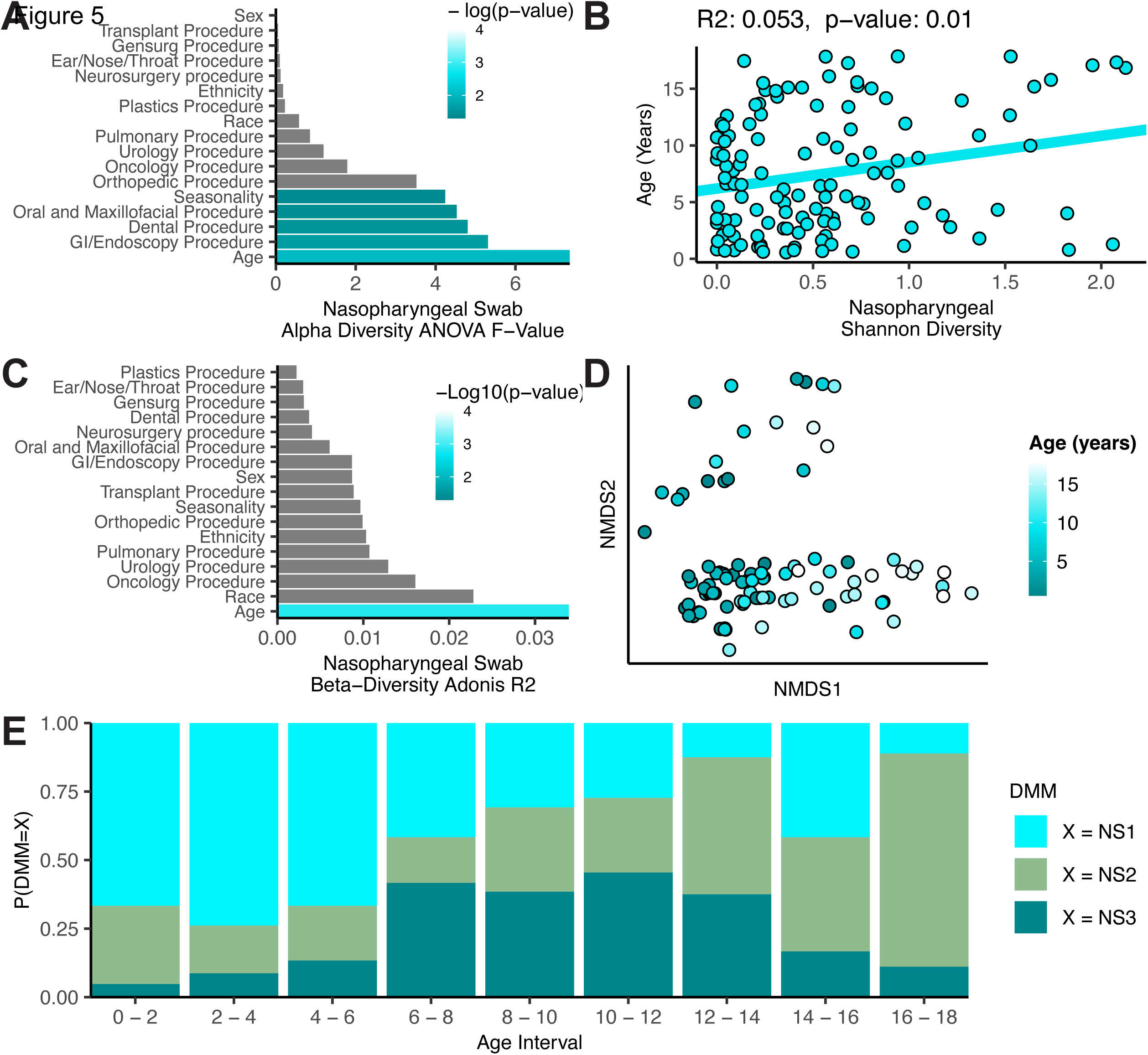
Nasopharyngeal microbiome composition correlates with age across childhood development. A) Results of an ANOVA comparing nasopharyngeal swab microbiome Shannon’s diversity against demographic features of the subjects. B) Pearson’s correlation between the subject age and nasopharyngeal microbiome Shannon’s diversity. C) Results of a PERMANOVA analysis comparing Unifrac distances between subjects against demographic features. D) Non-metric dimensional scaling plot rotated to show the influence of age on Unifrac distances between subjects. E) Bar plot depicting the distribution of nasopharyngeal swab clusters across age. All 124 nasopharyngeal swabs were used in each analysis.

By performing a similar analysis against the tracheal aspirate microbiome alpha diversities, we identified that subject-reported race and whether the subject underwent an ear/nose/or throat procedure explained a significant proportion of the variance in the data (Supplementary Figure S3C). We confirmed these findings by comparing the mean alpha diversities of tracheal aspirate microbiomes between subject-reported races (Supplementary Figure S3D) and between subjects undergoing ear/nose/throat procedure and those that were not (Supplementary Figure S3E). An analysis comparing the Unifrac distances between samples against demographics identified ethnicity, as well as whether the subject underwent an ear/nose/throat procedure or an orthopedic procedure each explained significant proportion of the variance in the Unifrac distances between tracheal aspirate microbiomes (Supplementary Figure S3F-G). While subject-reported race explained the greatest proportion of variance, it remained non-significant (adonis2 p-value > 0·07), likely because of multiple unevenly distributed groups.

Observing an interaction across ecological measures of the nasopharyngeal swab microbiomes and age, we next asked whether these interactions might be explained by the clusters predicted by Dirichlet multinomial modeling. Nasopharyngeal swab microbiome cluster was an important predictor of age in a generalized linear model (Chi-square test p-value < 0·0001), so we visualized this relationship by looking at the distribution of each cluster across age (Figure 5E). We observed that the *D. pigrum* dominated NS1 cluster was most common in early childhood from ages of 5 months to 6 years, but that the *S. aureus* dominated NS3 cluster became more common between the ages of 6 to 12. After 12 years of age, the mixed community cluster NS2 remained the most common. The trend from communities of lower diversity towards communities of higher diversity during childhood development, corroborates the findings observed by comparing age against the alpha or beta diversities of the nasopharyngeal swab microbiomes.

## Discussion

In this study we extracted, sequenced, and performed quality control on the nasopharyngeal swabs and tracheal aspirates of over 100 pediatric subjects. This unique cohort containing 85 subject-matched upper and lower respiratory tract samples granted us the opportunity to investigate the relationship between the upper and lower respiratory tract microbiomes, as well as explore the influence of age on the respiratory tract microbiota over the course of childhood development. Our study confirms major taxa of the upper and lower airway consistent with those found in previous literature, and we identified well-described interactions among the taxa colonizing the upper respiratory tract. These network relationships were captured by Dirichlet multinomial modeling as distinct clusters that share a modest correlation between the upper respiratory tract and lower respiratory tract. Importantly, we found that the clusters that define the upper respiratory tract share a strong relationship with age.

We found it surprising that the common metrics of ecologic richness did not correlate between the upper and lower respiratory tracts. Moreover, co-occurrence analysis did not identify taxa that might be commuting between the upper and lower respiratory tracts. We were only able to identify any relationship across these two sites by examining the correlation between microbiome clusters of the URT and those of the LRT. Interestingly, the strongest correlation between the highly mixed URT cluster and the *Bacillus*-dominated LRT cluster shared no taxa in common. There are a few possibilities that could explain the observed phenomenon. Firstly, the nasopharyngeal swab site and the tracheal aspirate are situated anatomically on either side of the oropharynx, a site with greater richness and biomass than the nasopharynx and trachea.^4^ The oropharyngeal microbiota may play an important role in contributing different taxa to the upper and lower respiratory tracts but serve as an environment where those taxa may interact. Alternatively, many host-extrinsic factors (e.g., seasonality) have been shown to affect the composition of the respiratory tract microbiota.^4^ While the taxa present in each anatomical location are distinct, the host extrinsic factors are constant within a given child, which might appear in our data as a correlation between separate upper and lower respiratory tract clusters.

When comparing demographic factors against measures of ecological diversity, we identified a strong effect of age on both the alpha and beta diversity of the nasopharyngeal swab samples. We find that the likelihood of an individual harboring a given URT cluster varied predictably with age. Previous studies of microbial succession of the URT have reported that healthy infants tend to be colonized by *Corynebacterium* spp. and *D. pigrum*.^28^ This was consistent with our finding that over the first few years of life, infants and toddlers were more likely to be colonized by a community that featured those microorganisms. Beyond early childhood, we saw a shift towards a *S. aureus*-dominated community between the ages of 6 to 12, and then a more diverse, mixed community through adolescence. Our findings suggest a stereotypic pattern of development for the microbiome of the upper respiratory tract.

Our findings are limited by the cross-sectional nature of the study, incomplete subject demographic information, and the technical limitations inherent to working with samples with low microbial biomass. While we have uncovered an interesting relationship between age and the composition of the upper respiratory tract microbiome, confirming these findings would require dedicated longitudinal study of potential shifts in microbiome composition in an individual over time. Additionally, the study was not designed to investigate the microbiome of the respiratory tract and important information that would strengthen our findings, such as vaccination status and medication use outside of antibiotics, was not collected. Several of the important external factors such as pollen levels, climate, and pollution were well controlled for in this population, as subjects were enrolled over a window of a few months and in a relatively constrained geographical area, but data about dietary, cultural, or socioeconomic differences that may be strongly correlated with subject race were not collected. Moreover, while the normal, heathy human airway is frequently exposed to respiratory viruses, this cohort, collected during the early COVID-19 epidemic, had no recent respiratory infections.

The microbiome data generated here was also sensitive to procedure type and it must be stated that our study population undergoing elective procedures does not represent an entirely healthy population. Procedure types were well distributed, but conditions requiring otolaryngological or maxillofacial procedures likely impact airway microbiome composition to an unknown extent. We did observe differences in alpha diversity associated with maxillofacial procedures in the URT and otolaryngological procedures in the LRT. We also observed that a portion of the variance in the Unifrac distances between LRT samples could be explained by otolaryngological procedures. Nevertheless this, we believe that these samples are an important window into the association between the respiratory microbiomes and human health and that the results generated here support further investigations in more focused studies.

Finally, nasopharyngeal swabs and tracheal aspirates represent low abundance microbial samples that are prone to contamination and bias. Our method for sequencing required an initial PCR amplification cycle to achieve sufficient DNA for adequate V4 16S rDNA sequencing, which is known to also increase the sensitivity toward contaminant sequences as well.^29^ Pre-amplification was performed with primers that target a larger region of the 16S rDNA sequence encompassing the V4 sequence to reduce the risk of generating contaminating V4 sequences. We also employed decontam to identify and remove likely contaminant sequences by comparing our samples against negative controls.^30^ While these methods are not guaranteed to completely resolve the challenges of low abundance sequences, we are encouraged by the findings in our data that match and support previous literature and our understanding of airway microbial ecology.

Our finding that the upper respiratory tract microbiota continues to develop through early life is striking. While the ecological succession of the microbial taxa in the upper respiratory tract has been described through the first two years of life, the continuation of a stereotypic pattern throughout childhood development implies a correlation with the physiological, immunological, and behavioral developments of the individual throughout life. A complete description of this pattern through childhood would provide a metric for understanding the degree and effects of dysbiosis induced by disturbance and treatment and could be used to predict an individual’s risk for disease. By understanding the natural patterns of ecological succession in the upper airways, we may begin shepherd development towards beneficial, health-associated patterns with vaccination or probiotics.

## Data Availability

All data produced in the present study are available upon reasonable request, or available through the European Nucleotide Archives (PRJEB65487).

## Contributors

AJH, ALR, ALK, ARJ conceived or designed the work. EEL, EHA, AMB, WRO, JL, LRY, and RMH conducted the study or collected the data. ALR and CPT extracted and processed collected samples. AJH processed the sequencing files, analyzed, and interpreted the data. All authors critically revised the article, verified the underlying data of the study, and had full access to the data in the study and final responsibility for the decision to submit for publication.

## Data Sharing

De-identified participant subject data and R code used for analysis will be made available after article publication upon written request. Raw V4 16S rDNA sequencing data have been deposited at the European Nucleotide Archive and are publicly available after publication under the project accession number PRJEB65487.

## References

1. Tony-Odigie, A., Wilke, L., Boutin, S., Dalpke, A. H. & Yi, B. Commensal Bacteria in the Cystic Fibrosis Airway Microbiome Reduce P. aeruginosa Induced Inflammation. Front Cell Infect Microbiol 12, 824101 (2022).

2. Teo, S. M. et al. Airway Microbiota Dynamics Uncover a Critical Window for Interplay of Pathogenic Bacteria and Allergy in Childhood Respiratory Disease. Cell Host Microbe 24, 341–352.e5 (2018).

3. van Meel, E. R., Jaddoe, V. W. V., Bønnelykke, K., de Jongste, J. C. & Duijts, L. The role of respiratory tract infections and the microbiome in the development of asthma: A narrative review. Pediatr Pulmonol 52, 1363–1370 (2017).

4. Man, W. H., de Steenhuijsen Piters, W. A. A. & Bogaert, D. The microbiota of the respiratory tract: gatekeeper to respiratory health. Nat Rev Microbiol 15, 259–270 (2017).

5. McCauley, K. E. et al. Seasonal airway microbiome and transcriptome interactions promote childhood asthma exacerbations. J Allergy Clin Immunol 150, 204–213 (2022).

6. Wilson, N. G., Hernandez-Leyva, A. & Kau, A. L. The ABCs of wheeze: Asthma and bacterial communities. PLoS Pathog 15, e1007645 (2019).

7. Proctor, D. M. & Relman, D. A. The Landscape Ecology and Microbiota of the Human Nose, Mouth, and Throat. Cell Host Microbe 21, 421–432 (2017).

8. Elgamal, Z., Singh, P. & Geraghty, P. The Upper Airway Microbiota, Environmental Exposures, Inflammation, and Disease. Medicina (Kaunas) 57, 823 (2021).

9. Yan, M. et al. Nasal microenvironments and interspecific interactions influence nasal microbiota complexity and S. aureus carriage. Cell Host Microbe 14, 631–640 (2013).

10. Dickson, R. P. et al. Spatial Variation in the Healthy Human Lung Microbiome and the Adapted Island Model of Lung Biogeography. Ann Am Thorac Soc 12, 821–830 (2015).

11. Pulvirenti, G. et al. Lower Airway Microbiota. Front Pediatr 7, 393 (2019).

12. Natalini, J. G., Singh, S. & Segal, L. N. The dynamic lung microbiome in health and disease. Nat Rev Microbiol 21, 222–235 (2023).

13. Charlson, E. S. et al. Topographical continuity of bacterial populations in the healthy human respiratory tract. Am J Respir Crit Care Med 184, 957–963 (2011).

14. Lin, E. E. et al. Concordance of Preprocedure Testing With Time-of-Surgery Testing for SARS-CoV-2 in Children. Pediatrics 147, e2020044289 (2021).

15. Lin, E. E. et al. Concordance of Upper and Lower Respiratory Tract Samples for SARS-CoV-2 in Pediatric Patients: Research Letter. Anesthesiology 134, 970–972 (2021).

16. Hazan, G. et al. Age-Dependent Reduction in Asthmatic Pathology through Reprogramming of Postviral Inflammatory Responses. J Immunol 208, 1467–1482 (2022).

17. Wilson, N. G. et al. The gut microbiota of people with asthma influences lung inflammation in gnotobiotic mice. iScience 26, 105991 (2023).

18. Davis, N. M., Proctor, D. M., Holmes, S. P., Relman, D. A. & Callahan, B. J. Simple statistical identification and removal of contaminant sequences in marker-gene and metagenomics data. Microbiome 6, 226 (2018).

19. Altschul, S. F., Gish, W., Miller, W., Myers, E. W. & Lipman, D. J. Basic local alignment search tool. J Mol Biol 215, 403–410 (1990).

20. Brugger, S. D. et al. Dolosigranulum pigrum Cooperation and Competition in Human Nasal Microbiota. mSphere 5, e00852–20 (2020).

21. Santacroce, L. et al. The Human Respiratory System and its Microbiome at a Glimpse. Biology (Basel) 9, 318 (2020).

22. Lasek, R. et al. Genome Structure of the Opportunistic Pathogen Paracoccus yeei (Alphaproteobacteria) and Identification of Putative Virulence Factors. Front Microbiol 9, 2553 (2018).

23. Daneshvar, M. I. et al. Paracoccus yeeii sp. nov. (formerly CDC group EO-2), a novel bacterial species associated with human infection. J Clin Microbiol 41, 1289–1294 (2003).

24. Kotiranta, A., Lounatmaa, K. & Haapasalo, M. Epidemiology and pathogenesis of Bacillus cereus infections. Microbes Infect 2, 189–198 (2000).

25. Du, T., Lei, A., Zhang, N. & Zhu, C. The Beneficial Role of Probiotic Lactobacillus in Respiratory Diseases. Front Immunol 13, 908010 (2022).

26. Fangous, M.-S. et al. Prevalence and dynamics of Lactobacillus sp. in the lower respiratory tract of patients with cystic fibrosis. Res Microbiol 169, 222–226 (2018).

27. Mathieu, E. et al. Paradigms of Lung Microbiota Functions in Health and Disease, Particularly, in Asthma. Front Physiol 9, 1168 (2018).

28. Schenck, L. P., Surette, M. G. & Bowdish, D. M. E. Composition and immunological significance of the upper respiratory tract microbiota. FEBS Lett 590, 3705–3720 (2016).

29. Schenck, L. P. et al. Nasal Tissue Extraction Is Essential for Characterization of the Murine Upper Respiratory Tract Microbiota. mSphere 5, e00562–20 (2020).

30. Pérez-Cobas, A. E., Rodríguez-Beltrán, J., Baquero, F. & Coque, T. M. Ecology of the respiratory tract microbiome. Trends Microbiol 31, 972–984 (2023).

