## Supplementary material for "Developmental progression of the nasopharyngeal microbiome during childhood and association with the lower airway microbiome": tansea_supp_material_submit_v4.docx

**Decontamination and filtering of low microbial abundance samples from nasopharyngeal swabs and tracheal aspirates**

To understand the relationship between the upper and lower airway microbiota, we profiled the microbial composition of the nasopharyngeal swabs and tracheal aspirates collected from children under the age of 18 years using V4 16S rDNA sequencing (see Methods).1 Both these sample types are of low microbial abundance and present challenges for analysis because contaminants from reagents or the environment make up a proportionally larger fraction of reads and artificially inflate classic metrics of microbial diversity such as alpha diversity.2 Several methods have been proposed to identify and remove contaminating sequences from metagenomic data generated from low microbial abundance samples.3 Here we filtered contaminant sequences from amplicon sequence variants (ASVs) generated by DADA2 with the R package decontam, which models the likelihood of a sequence being a contaminant based on its rate of detection in negative control samples (prevalence model) and the relationship between its abundance and the original DNA concentration of the sample (frequency model).4

Thresholds for contaminant prediction from the frequency model (Supplementary Figure S1A,B) and the prevalence model (Supplementary Figure S1C,D) were estimated separately for nasopharyngeal swabs and tracheal aspirates based on the distributions of scores predicted for each model. ASVs classified as a contaminant by either model were removed from the respective set of samples. We were able to extract DNA from all 183 nasopharyngeal swabs. After decontamination 159 samples still had greater than 1000 reads and were retained. Only 166 of the 183 tracheal aspirates extracted had sufficient DNA to sequence. Of the 166 tracheal aspirates extracted and sequenced, 131 still had greater than 1000 reads after decontamination and were retained.

After decontamination, we used PERMANOVA to identify whether biological differences were still captured by the data. As expected, sample type (nasopharyngeal swab compared to tracheal aspirate) explained a significant proportion of the variation (adonis2 R2: 15·1%, p-value < 0·001). However, there was a strong association between extraction round and apparent taxonomic composition (adonis2 R2: 7·02%, p-value: 0·002) (Supplementary Figure S1E). The inconsistent extraction rounds were identified using a post-hoc test (Supplementary Figure S1F) and those samples were removed from further analysis. This reduced the overall influence of extraction round on the data (adonis2 R2 = 5·3%, p-value: 0·098) without significantly impacting the expected biological signal (adonis2 R2: 15·7%, p-value < 0·001). Ultimately, data from 124 nasopharyngeal swabs and 98 tracheal aspirates were included in the final analysis (Supplementary Figure S1G). Of these, 85 nasopharyngeal swabs and tracheal aspirates represented paired samples collected from the same subject.

The demographics of the remaining subjects are summarized in Table 1. Between the 124 nasopharyngeal swabs and the 98 tracheal aspirates, 85 pairs of samples are subject-matched. The demographics are largely well-matched and unremarkable, apart from intervention type (Supplementary Table 1). Samples from subjects that underwent pulmonary bronchoscopic procedures and/or gastrointestinal endoscopy are only represented among the nasopharyngeal swabs. The matching tracheal aspirate samples were filtered out during the decontamination phase. Regardless, to our knowledge, these data represent one of the largest reported cohorts of matched upper and lower respiratory tract samples in children.

**Supplementary Figure S1**

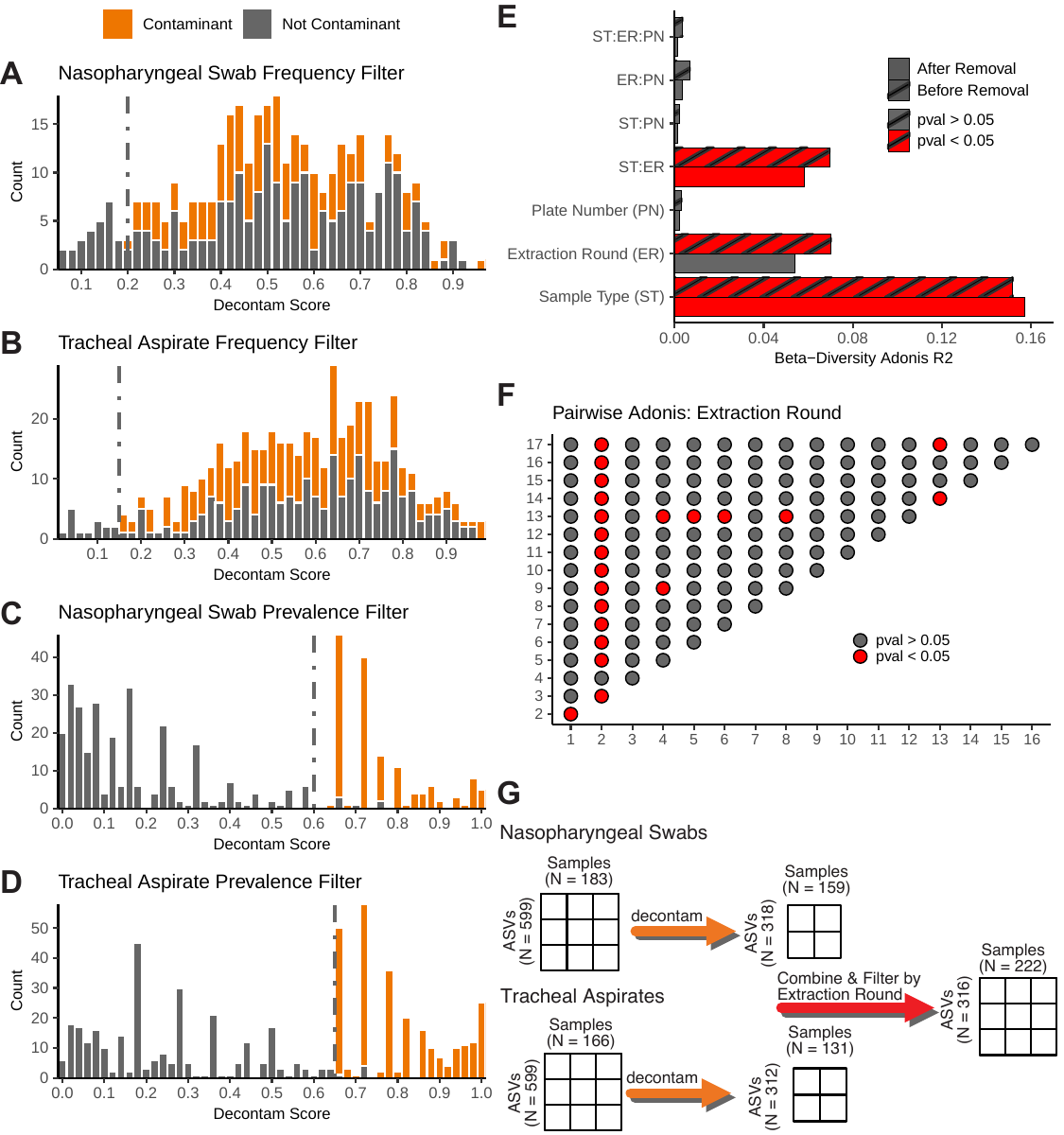

**Supplementary Figure S1: Decontamination and filtering of V4 16S rDNA sequencing of 184 nasopharyngeal swabs and 166 tracheal aspirates.** A - B) Histogram of scores generated by decontam fitting the nasopharyngeal swabs and tracheal aspirates to the frequency model for contamination. C-D) Histogram of scores generated by decontam fitting the nasopharyngeal swabs and tracheal aspirates against the prevalence model for contamination. For histograms A-D, dashed, gray vertical lines indicate user-selected thresholds. Gray bars represent counts of ASVs identified as contamination within either nasopharyngeal swabs or tracheal aspirates by either filter. Orange bars represent counts of ASVs that were retained. E) Bar plot depicting the results of the PERMANOVA comparing nasopharyngeal and tracheal samples after filtering with decontam against batch variables and sample type. Striped patterns represent PERMANOVA results before removal of highly discriminatory extraction rounds. F) Pairwise-adonis analysis was used as a post-hoc test to identify extraction rounds that were distinct from all others. G) Diagram depicting the results of decontamination and filtering.

**Supplementary Figure S2**

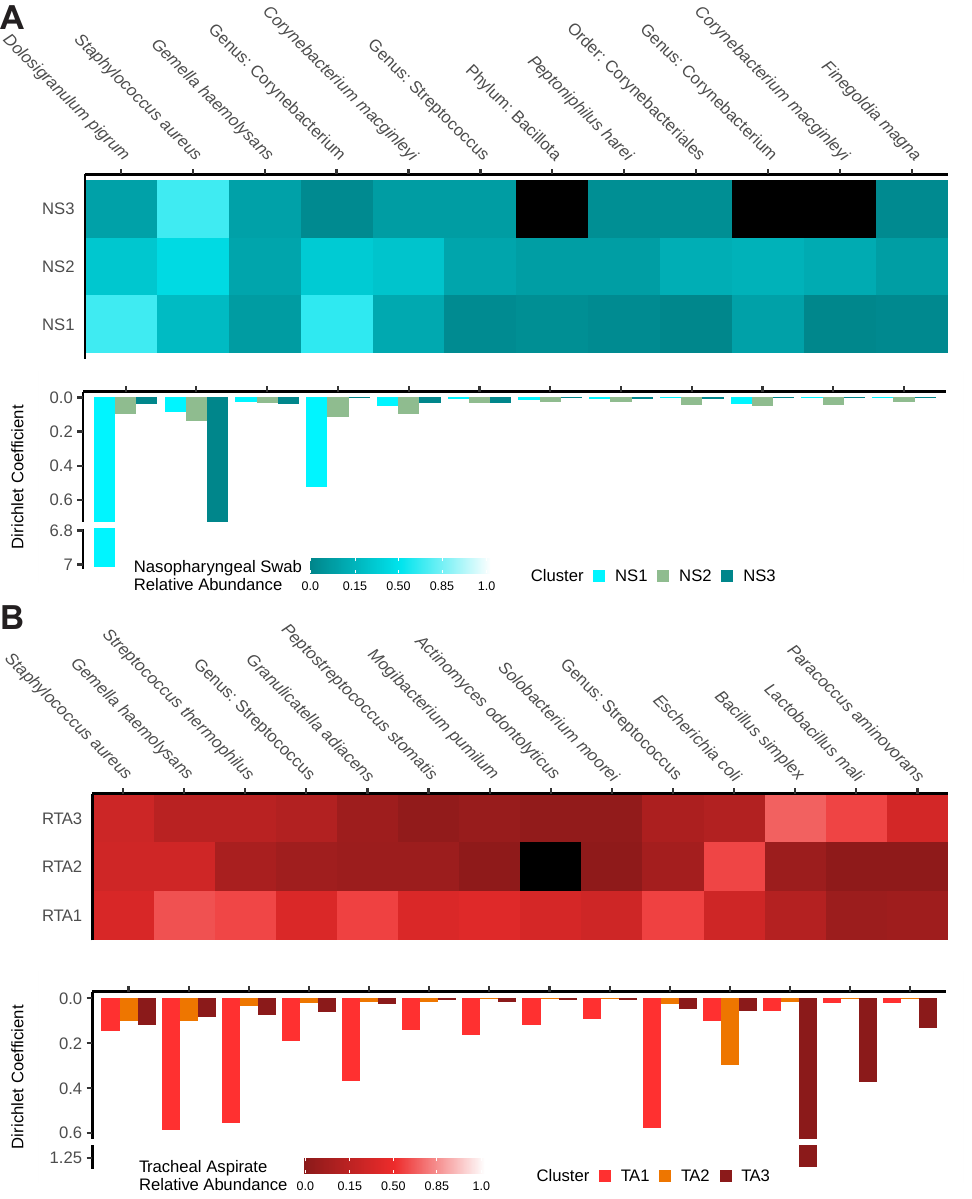

**Supplementary Figure S2: Dirichlet multinomial modeling predicts three distinct community clusters each in the nasopharyngeal swabs and the tracheal aspirates.** A-B) Heatmaps and bar plots describing the results of Dirichlet multinomial modeling of the nasopharyngeal microbiomes (A) and tracheal aspirate microbiomes (B). The heatmaps describe the frequency we detected a particular taxon in subjects separated by DMM cluster. The bar plots describe, for the relevant taxon and for each cluster, the Dirichlet multinomial model coefficient. Nasopharyngeal swabs are divided into three clusters: NS1 (N = 60), NS2 (N = 38), and NS3 (N = 26). Tracheal aspirates are divided into three clusters: TA1 (N = 50), TA2 (N = 29), and TA3 (N = 19).

**Supplementary Figure S3**

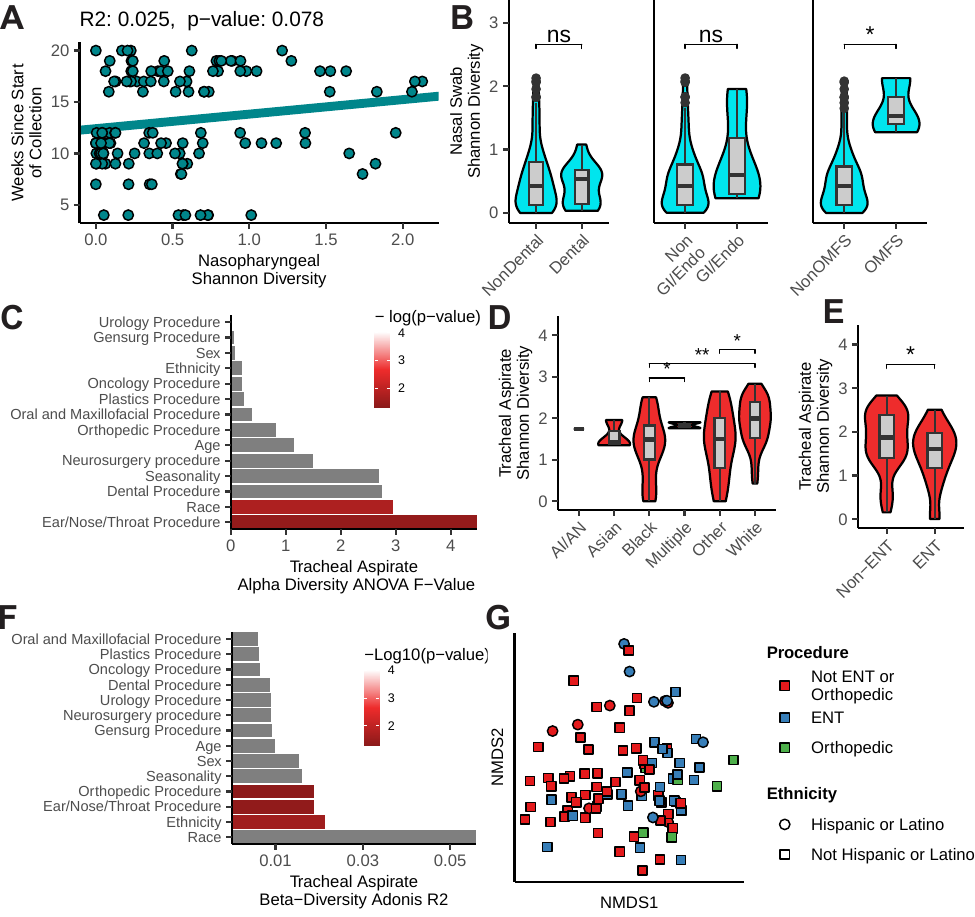

**Supplementary Figure S3: Comparing upper and lower airway microbiome features against subject demographics.** A) Pearson’s correlation between seasonality of sample collection and nasopharyngeal swab Shannon’s diversity (N = 124). B) Univariate (t-test) comparison of nasopharyngeal swab Shannon’s diversity between subjects receiving dental (N = 13) vs. non-dental procedures (N = 111), subject receiving gastrointestinal/endoscopic (GI/Endo N = 7) vs. non-gastrointestinal/endoscopic procedures (N = 117), and subjects receiving oral/maxillofacial procedures (OMFS, N = 3) vs. non-oral/maxillofacial procedures (N = 121). C) Results of an ANOVA comparing tracheal aspirate microbiome Shannon’s diversity against demographic features of the subjects (N = 98). D) Univariate analysis (t-tests) comparing tracheal aspirate Shannon’s diversity across subject self-reported race (American Indian/Alaska Native N = 1, Asian N = 3, Black N = 21, Multiple N = 2, Other N = 17, White N = 54). E) Univariate analysis (t-test) comparing tracheal aspirate Shannon’s diversity between subjects undergoing ear, nose, and throat procedures (ENT N = 34) against subjects not undergoing ear, nose, and throat procedures (n = 64). F) Results of a PERMANOVA comparing Unifrac distances between tracheal aspirate microbiomes and subject’s demographic features (N = 98). G) Non-metric dimensional scaling comparing Unifrac distances between tracheal aspirate microbiomes and subject demographic features. Subjects undergoing ear/nose/throat procedures are colored blue (N = 34). Subjects undergoing orthopedic procedures are colored green (N = 6). Subjects undergoing procedures that are neither ear/nose/throat or orthopedic are colored red (N = 58). Subjects reporting Hispanic or Latino ethnicity are represented by circular points (N = 14) and subjects reporting not Hispanic or Latino are represented by square points (N = 84).

**Supplementary Table 1: Summary of statistical tests performed within the study**

| **Comparison** | **Test** | **Feature** | **Estimate** | **p-value** |
| --- | --- | --- | --- | --- |
| RTA vs. NSWB | t.test | Age | 0·62302 | 0·5339 |
| RTA vs. NSWB | t.test | Seasonality | -0·2169 | 0·8285 |
| RTA vs. NSWB | chisq | Sex | 0·044966 | 0·8321 |
| RTA vs. NSWB | chisq | Any chronic respiratory disease | 0·17042 | 0·6797 |
| RTA vs. NSWB | chisq | Any chronic disease | 0·2338 | 0·6287 |
| RTA vs. NSWB | chisq | Born Premature | 0 | 1 |
| RTA vs. NSWB | chisq | Race | 1·0069 | 0·962 |
| RTA vs. NSWB | chisq | Ethnicity | 0 | 1 |
| RTA vs. NSWB | chisq | General Procedure | 0·12269 | 0·7261 |
| RTA vs. NSWB | chisq | Ear/Nose/Throat Procedure | 0·13541 | 0·7138 |
| RTA vs. NSWB | chisq | Urologic Procedure | 0·68787 | 0·4069 |
| RTA vs. NSWB | chisq | Orthopedic Procedure | 0 | 1 |
| RTA vs. NSWB | chisq | NeuroProcedure | 0 | 1 |
| RTA vs. NSWB | chisq | Dental Procedure | 0 | 1 |
| **RTA vs. NSWB** | **chisq** | **Pulmonary Procedure*** | **4·8332** | **0·02792** |
| **RTA vs. NSWB** | **chisq** | **Gastrointestinal Endoscopy*** | **4·0133** | **0·04514** |
| RTA vs. NSWB | chisq | Plastics Procedure | 0 | 1 |
| RTA vs. NSWB | chisq | Oral and Maxillofacial Procedure | 0 | 1 |
| **NP DMM** | **Anova** | **Age***** | **9·898** | **0·000104** |
| ***NS2 v. NS1**** | ***TukeyHSD*** | ***Age*** | ***4·1158832*** | ***0·0003164*** |
| ***NS3 v. NS1**** | ***TukeyHSD*** | ***Age*** | ***3·719294*** | ***0·005046*** |
| *NS3 v. NS2* | *TukeyHSD* | *Age* | *-0·3965892* | *0·9470482* |
| NP DMM | Anova | Seasonality | 2·823 | 0·0633 |
| NP DMM | chisq | Sex | 0·64927 | 0·7228 |
| NP DMM | chisq | Any chronic respiratory disease | 0·53549 | 0·7651 |
| NP DMM | chisq | Any chronic disease | 5·7835 | 0·05548 |
| NP DMM | chisq | Born Premature | 0·73338 | 0·693 |
| NP DMM | chisq | Race | 7·943 | 0·6344 |
| NP DMM | chisq | Ethnicity | 3·896 | 0·1426 |
| NP DMM | chisq | General Procedure | 0·19783 | 0·9058 |
| NP DMM | chisq | Ear/Nose/Throat Procedure | 0·82093 | 0·6633 |
| NP DMM | chisq | Urologic Procedure | 4·0537 | 0·1317 |
| **NP DMM** | **chisq** | **Orthopedic Procedure***** | **9·4002** | **0·009094** |
| NP DMM | chisq | NeuroProcedure | 0·33501 | 0·8458 |
| NP DMM | chisq | Dental Procedure | 1·6535 | 0·4375 |
| NP DMM | chisq | Pulmonary Procedure | 4·4933 | 0·1058 |
| NP DMM | chisq | Gastrointestinal Endoscopy | 1·1676 | 0·5578 |
| NP DMM | chisq | Plastics Procedure | 2·335 | 0·3111 |
| **NP DMM** | **chisq** | **Oral and Maxillofacial Procedure*** | **6·9578** | **0·03084** |
| RTA DMM | Anova | Age | 1·036 | 0·359 |
| RTA DMM | Anova | Seasonality | 0·131 | 0·877 |
| RTA DMM | chisq | Sex | 1·6198 | 0·4449 |
| RTA DMM | chisq | Any chronic respiratory disease | 1·1763 | 0·5554 |
| RTA DMM | chisq | Any chronic disease | 0·019591 | 0·9903 |
| RTA DMM | chisq | Born Premature | 1·0032 | 0·6055 |
| RTA DMM | chisq | Race | 16·612 | 0·08341 |
| **RTA DMM** | **chisq** | **Ethnicity*** | **7·6506** | **0·02181** |
| RTA DMM | chisq | General Procedure | 0·97266 | 0·6149 |
| **RTA DMM** | **chisq** | **Ear/Nose/Throat Procedure**** | **11·531** | **0·003134** |
| RTA DMM | chisq | Urologic Procedure | 1·286 | 0·5257 |
| RTA DMM | chisq | Orthopedic Procedure | 1·0632 | 0·5877 |
| RTA DMM | chisq | NeuroProcedure | 1·2265 | 0·5416 |
| RTA DMM | chisq | Dental Procedure | 5·1307 | 0·07689 |
| RTA DMM | chisq | Pulmonary Procedure | NA | NA |
| RTA DMM | chisq | Gastrointestinal Endoscopy | NA | NA |
| RTA DMM | chisq | Plastics Procedure | 0·098314 | 0·952 |
| RTA DMM | chisq | Oral and Maxillofacial Procedure | 0·76313 | 0·6828 |

**References**

1.	Yu G, Fadrosh D, Goedert JJ, Ravel J, Goldstein AM. Nested PCR Biases in Interpreting Microbial Community Structure in 16S rRNA Gene Sequence Datasets. PLoS One. 2015;10(7):e0132253.

2.	de Goffau MC, Lager S, Salter SJ, Wagner J, Kronbichler A, Charnock-Jones DS, et al. Recognizing the reagent microbiome. Nat Microbiol. 2018 Aug;3(8):851–3.

3.	Karstens L, Asquith M, Davin S, Fair D, Gregory WT, Wolfe AJ, et al. Controlling for Contaminants in Low-Biomass 16S rRNA Gene Sequencing Experiments. mSystems. 2019 Jun 4;4(4):e00290-19.

4.	Davis NM, Proctor DM, Holmes SP, Relman DA, Callahan BJ. Simple statistical identification and removal of contaminant sequences in marker-gene and metagenomics data. Microbiome. 2018 Dec 17;6(1):226.
